# Transcatheter palliation with pulmonary artery flow restrictors in neonates with congenital heart disease: feasibility, outcomes, and comparison with a historical hybrid Stage 1 cohort

**DOI:** 10.1101/2023.05.15.23290017

**Authors:** Francesca Sperotto, Nora Lang, Meena Nathan, Aditya Kaza, David M Hoganson, Eleonore Valencia, Catherine K Allan, Eduardo M Da Cruz, Pedro J Del Nido, Sitaram M Emani, Christopher Baird, Nicola Maschietto

**Affiliations:** Department of Cardiology, Boston Children’s Hospital, Harvard Medical School, Boston, USA; Department of Pediatric Cardiology, University Heart & Vascular Center Hamburg, University Medical Center Hamburg-Eppendorf, Hamburg, Germany; Department of Cardiac Surgery, Boston Children’s Hospital, Harvard Medical School, Boston, USA

## Abstract

**Background:** Neonates with complex congenital heart disease (CHD) and pulmonary overcirculation have been historically treated surgically. However, sub-cohorts of patients may benefit from less invasive procedures. Transcatheter palliation with pulmonary flow restrictors (PFRs) may represent a compelling alternative, but data are limited.

**Methods:** We present our experience of palliation with PFRs in neonates with CHD and pulmonary overcirculation, including procedural feasibility, technical details, and patient-level outcomes. We then compared our sub-cohort of high-risk single ventricle (SV) neonates palliated with PFRs with a historical cohort of high-risk SV neonates palliated with a hybrid Stage 1. Cox regression was used to evaluate the association between palliation strategy and all-cause mortality risk at 6 months.

**Results:** From 2021 to 2023, 17 patients (median age 4 days, interquartile range [IQR] 2-8); median weight 2.51 kilograms [IQR 2.09-3.26]) underwent a PFR procedure; 15 (88%) had SV physiology; 15 (88%) were considered high-risk surgical candidates. All the procedures were technically successful. At a median follow-up of 5.3 months (IQR 1.9-9.6), 13 patients (76%) were either successfully bridged to surgery (n=10, 59%) or are awaiting surgery (n=3, 17%). Patients underwent the target surgery after a median of 2.6 months (IQR 1.2-3.4) from the PFR procedure (median weight 4.6 kilograms [IQR 3.2-5.4]). Their pulmonary arteries were found to have grown adequately for age. All PFR devices were easily removed without the need for arterioplasty. The all-cause mortality rate before target surgery was 24% (n=4). Compared to a historical cohort of high-risk SV neonates palliated with a hybrid Stage 1 (n=23), after adjustment for main confounding (age, weight, presence of intact atrial septum or severely restrictive patent foramen ovale, and presence of left ventricle to coronary fistulae), the PFR procedure was associated with a significantly lower all-cause 6-month mortality risk (adjusted Hazard Ratio=0.30 [95% CI 0.10- 0.93]).

**Conclusions:** Transcatheter PFR palliation in high-risk neonates with CHD is feasible, safe, and may represent an effective alternative strategy to bridge such high-risk neonates to surgical palliation, complete repair, or transplant while allowing for clinical stabilization and somatic growth.

**Clinical perspectives:** *What is new?:*

- Transcatheter PFR palliation in high-risk neonates with congenital heart disease is feasible, safe, and effective in reducing pulmonary blood flow and allow for clinical stabilization and growing.
- PFR devices can be easily removed both at cardiac catheterization or surgery with no need for pulmonary arterioplasty, and pulmonary artery grow adequately over time.
- Compared to a historical cohort of high-risk single ventricle neonates palliated with a hybrid Stage-1, after adjustment for main confounding, the PFR palliation was shown to be associated with a significantly lower 6-month all-cause mortality risk.

*What are the clinical implications?:*

- Transcatheter PFR palliation in high-risk neonates may represent an effective alternative strategy to bridge such high-risk neonates to surgical palliation, complete repair, or transplant while avoiding a surgical procedure and allowing for clinical stabilization and somatic growth.

## Introduction

Neonates with complex congenital heart disease (CHD) and excessive pulmonary blood flow (PBF), such as those with single ventricle (SV) physiology and unbalanced circulations, are at high risk of adverse outcomes including pulmonary edema, low cardiac output, multi-organ failure, and cardiovascular collapse^1, 2^. Historically, patients with pulmonary overcirculation have been treated surgically, with various procedures based on the underlying anatomy. SV neonates with ductal-dependent systemic circulation, such as those with hypoplastic left heart syndrome (HLHS), typically undergo either a Norwood Stage 1 operation or a hybrid Stage 1 procedure. Both procedures aim to establish adequate mixing at the atrial level, unobstructed systemic flow, and controlled PBF^1, 3^. Neonates with other CHD and pulmonary overcirculation either undergo primary repair or palliation with a pulmonary artery (PA) band to control excessive PBF with subsequent complete repair^4^. These surgical procedures entail an open chest operation with or without cardio-pulmonary bypass in the neonatal period and are associated with non-negligible risks of morbidity and mortality^4, 5^. Importantly, sub cohorts of neonates, such as those with multiple risk factors like prematurity, low birth weight, significant comorbidities, ventricular dysfunction, or severe atrioventricular valve regurgitation (AVVR), may initially benefit from less invasive catheter-based approaches.

To date, reported experience regarding transcatheter pulmonary flow restrictors (PFRs) to palliate neonates with CHD is limited to two small case series and a few case reports^6–9^. We report our experience with PFRs in a cohort of neonates who were deemed to be high-risk surgical candidates. We describe the procedure feasibility, technical details, patient-level outcomes, and compared a sub cohort of high-risk SV patients palliated with a full transcatheter approach to a similar historical cohort that underwent a hybrid Stage 1 palliation.

## Methods

### Setting, design, and population

We performed a retrospective review of all consecutive patients who underwent an attempted transcatheter PFR placement from December 2021 to March 2023 at our center. Consent was obtained before the procedure. Additionally, the sub cohort of high-risk SV patients who underwent transcatheter PFR placement were compared to a historical cohort of SV neonates palliated with a hybrid Stage 1 procedure between January 2012 and March 2023. The Institutional Review Board at Boston Children’s Hospital approved the study with a waiver for informed consent (IRB-P00044617).

### Data collection, definitions, and outcome measures

Demographics, anatomical details, clinical characteristics, and patient-level outcomes were collected from patients’ charts and the institutional cardiac catheterization and surgical databases. For the PFR cohort, procedural and hemodynamic data were also collected. A procedure was considered technically successful when the patient left the catheterization laboratory with a PFR device implanted in each pulmonary artery (**Figure 1**) and clinical evidence of PBF restriction. Patients were diagnosed with left ventricle (LV) to coronary fistulae when anomalous connections between the LV cavity and the epicardial coronary vessels were angiographically confirmed. The vasoactive inotropic score was used to assess the level of pharmacologic hemodynamic support after the procedure^10^. PFR migration was defined as *acute* when it occurred immediately after the device release, or *late* when recognized in follow-up. PFR migration was also characterized as *partial* when the migrated device was still able to provide PBF restriction to all segments of the lungs, or *complete* when one or more segments had unrestricted PBF. Left atrial (LA) hypertension was defined as a measured LA pressure >10 mmHg.

**Figure 1.**
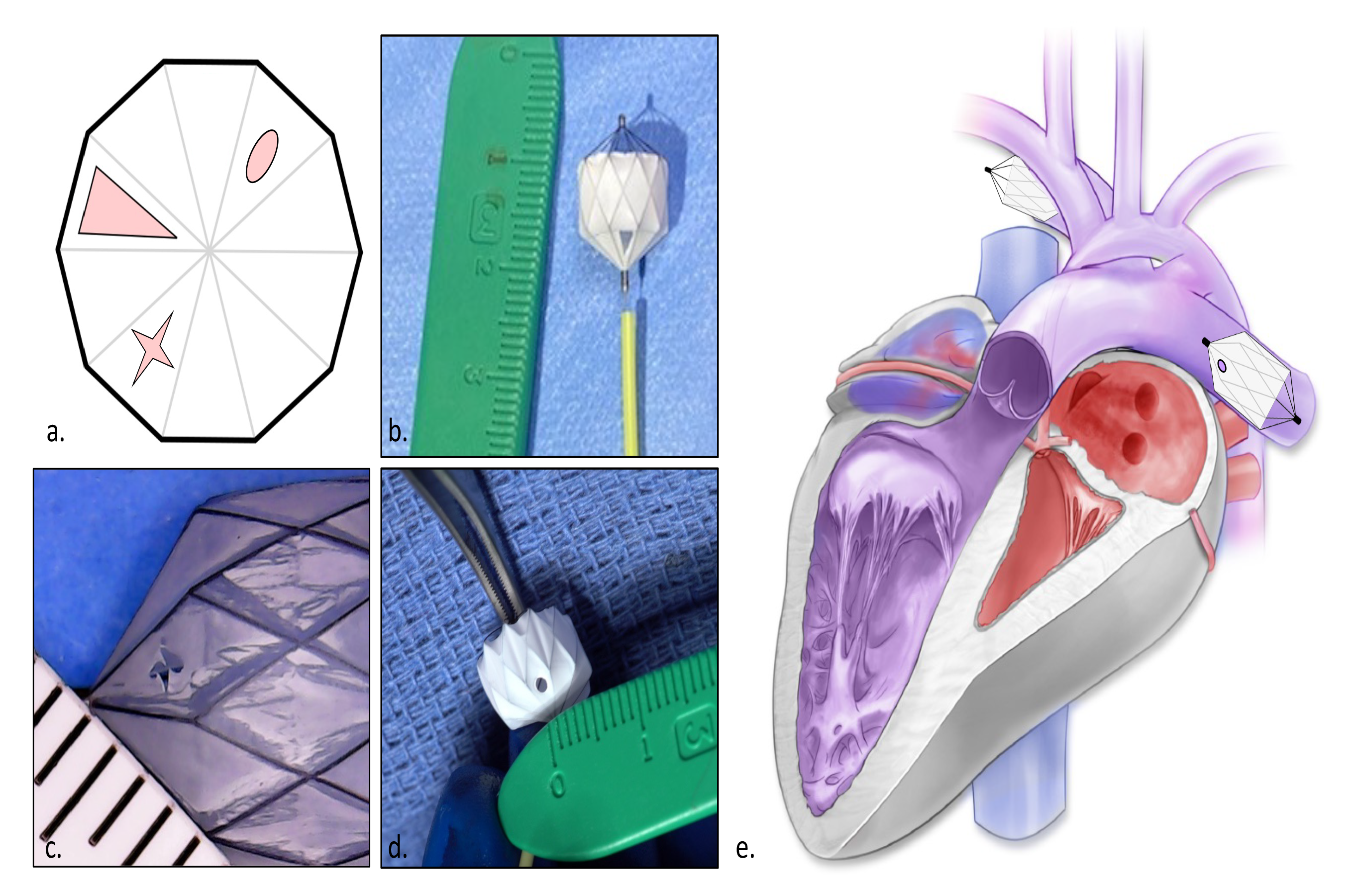
Pulmonary flow restrictors preparation and graphical example of their placement in a single ventricle heart. **a.** Graphical representation of different types of fenestrations. **b.** Initial 2-mm triangular section fenestration. **c.** Subsequent ∼1×1mm cross-shaped slit fenestration. **d.** Final 1-1.5 mm fenestration strategy. **e.** Graphical representation of pulmonary flow restrictor placement in the pulmonary arteries of an hypoplastic left heart.

Patient-level outcomes included procedural outcomes (successful or unsuccessful deployment and safety details) as well as 3-month and 6-month all-cause mortalities. For comparison between the PFR and hybrid cohorts, our primary outcome measure was mortality at 6 months; secondary outcome measures were mortality at 3 months and length of hospitalization.

### Procedural technique

All procedures were conducted under general anesthesia. A custom-made fenestrated Micro Vascular Plug (MVP; Medtronic, Minneapolis, MN, USA) was implanted as a PFR in each PA following a previously described technique^6–9, 11^. To appropriately control PBF, the fenestration technique was modified over time, as shown in **Figure 1** and reported in detail in the **Supplemental Methods**. To minimize the risk of distal device embolization, most of the MVPs were intentionally oversized by one size beyond the manufacturer’s recommendations. However, despite this precaution, device migrations still occurred, prompting us to modify the technique for subsequent patients. Beginning with patient #7, we were able to prevent right PA device migration by lodging the distal tip of the MVP inside the take-off of the right upper PA. The same technique could not be used on the left side because of the short anatomical distance between the upper lobe and the branch PA bifurcation. The technique for transcatheter PFR retrieval is described in the **Supplemental Methods**. After the procedure, patients received an infusion of heparin for 24 hours, targeting an activated partial thromboplastin time between 60 to 80 seconds, and started on dual antiplatelet therapy with aspirin and clopidogrel.

### Statistical Analysis

Descriptive data are summarized using frequencies and percentages for categorical variables, medians and interquartile ranges (IQR) for continuous variables. Changes in continuous variables over time were tested using the non-parametric Wilcoxon signed test. Comparisons of data between independent groups were performed using the Wilcoxon sum rank, Chi-squared, and Fisher’s exact tests as appropriate. Kaplan-Meier curves were used to estimate survival probabilities at 3 and 6 months. Cox regression was used to test the unadjusted and adjusted association between the palliation technique and time to all-cause mortality at 6-months. Proportional hazards assumptions were checked by comparing the log-log curve versus log-time. The multivariable model was adjusted for factors that were clinically deemed to be important confounders, i.e. age and weight at procedure, presence of an intact atrial septum (IAS) or severely restrictive patent foramen ovale (PFO), and presence of LV to coronary fistulae. Age and weight were tested for collinearity; since collinearity was not proven (Spearman Rho=0.064), they were both entered in the model. Number of events per variable were relaxed to n=5-9 based on Vietinghoff et al.’s study^12^. To avoid model overfitting and given their frequent clinical association, IAS or severely restrictive PFO and LV to coronary fistulae were grouped together as a single variable. Mortality risks were reported as hazard ratios (HRs) and 95% confidence intervals (CIs). All statistical analyses were performed using R Statistics (version 3.6.2; R Core Team, R Foundation for Statistical Computing, Vienna, Austria). Statistical significance was set at a two-sided *p*-value <0.05.

## Results

### Population

From December 2021 to March 2023, 17 patients (70% males) underwent an attempted transcatheter PFR placement. Most patients had SV physiology (n=15, 88%) and/or were high-risk surgical candidates (n=15, 88%). Among the 15 patients with SV physiology, the PFR procedure was intended as a bridge to surgical SV palliation in 8, bi-ventricular (BiV) repair in 5, and transplant in 2. In the 2 patients with a BiV physiology, the PFR procedure was intended as a bridge to complete surgical repair.

Patients’ demographic, anatomic, and clinical characteristics are summarized in **Table 1 and Supplemental Table 1**. The median age and weight at the time of procedure were 4 days (IQR 2-8) and 2.51 kg (IQR 2.09-3.26), respectively, and the smallest patient weighed 1.4 kilograms. The most common diagnosis was HLHS (n=11, 65%), with 4 patients having LV to coronary fistulae, 3 having an IAS requiring emergent atrial septal stenting immediately after delivery, 5 having a severely restrictive PFO, 1 having severe AVVR and severe ventricular dysfunction, and 1 having suspected pulmonary vein stenosis.

**Table 1.** Demographic, anatomic, and clinical characteristics of patients undergoing transcatheter pulmonary flow restrictors placement.

| **Variable** | **Total**<br>**(n=17)** |
| --- | --- |
| Age at PFR procedure, days | 4 (2-8) |
| Weight at PFR procedure, kg | 2.51 (2.09-3.26) |
| Gender, male | 12 (70.5) |
| Race |  |
| White | 10 (58.9) |
| Black/African American | 2 (11.7) |
| Hispanic | 3 (17.7) |
| Not available | 2 (11.7) |
| Gestational age at birth, weeks | 37.0 (33.8 -38.7) |
| Birth weight, kg | 2.42 (1.77-3.17) |
| Cardiac anatomic diagnosis |  |
| HLHS | 11 (64.7) |
| DILV | 1 (5.9) |
| DORV, hypoplastic LV | 2 (11.7) |
| VSD | 1 (5.9) |
| AP window, VSD | 1 (5.9) |
| LV aneurysm | 1 (5.9) |
| Circulation physiology at PFR procedure |  |
| Single ventricle | 15 (88.2) |
| Bi-ventricular | 2 (11.8) |
| Interventions before PFR procedure |  |
| Fetal balloon aortic valvuloplasty | 1 (5.9) |
| Atrial septum intervention | 3 (17.7) |
| Surgical PA banding | 1 (5.9) |
| Major baseline comorbid conditions |  |
| Prematurity (<37 weeks of gestation) | 9 (52.9) |
| Genetic syndrome and/or other congenital anomalies | 4 (23.5) |
| History of IUGR | 3 (17.7) |
| SGA (weight <10 %ile) | 3 (17.7) |
| Fetal hydrops | 1 (5.9) |
| Severe lung disease | 3 (17.7) |
| Severe infection, sepsis or septic shock | 2 (11.7) |
| NEC | 2 (11.7) |
| Neurologic disease | 2 (11.7) |
| Invasive MV at PFR procedure | 7 (41.1) |
| Inotropic support at PFR procedure | 8 (47.0) |
| Ventricular dysfunction at PFR procedure |  |
| Normal/Mild | 14 (82.3) |
| Moderate | 1 (5.9) |
| Severe | 2 (11.7) |
| Systemic AVVR at PFR procedure |  |
| None/Mild | 15 (88.2) |
| Moderate | 1 (5.9) |
| Severe | 1 (5.9) |

\*Prematurity was defined as a gestational age <37 weeks of gestation. Small for gestational age (SGA) was defined as a birth weight <10 percentile of normative data.
Continuous data are presented as median (Inter Quartile Range-IQR= 25% and 75%), categorical data are presented as frequency (%). Abbreviations: AP: aortopulmonary; AVVR: atrioventricular valve regurgitation; DILV: double inlet left ventricle; DORV: double outlet right ventricle; HLHS: hypoplastic left heart syndrome; IUGR: intrauterine growth retardation; Kg: kilogram; LV: left ventricle; MV: mechanical ventilation; NEC: necrotizing enterocolitis; PA: pulmonary artery; PFR: Pulmonary Flow Restrictor; SGA: small for gestational age; VSD: ventricular septal defect.

In terms of comorbidities, 53% (n=9) of patients were premature with the most premature neonate born at 26 weeks of gestation, 23% (n=4) had a genetic syndrome or congenital anomalies, 18% (n=3) had severe lung disease, 12% (n=2) had necrotizing enterocolitis, and 12% (n=2) had a neurologic disease. At the time of PFR procedure, 41% (n=7) were supported with mechanical ventilation, 47% (n=8) with inotropic medications; 12% (n=2) had more than mild SV systolic dysfunction, and 12% (n=2) more than mild systemic AVVR. One patient had undergone a prior surgical operation (arch reconstruction and PA band) before the PFR procedure.

### Procedural details and outcomes

All attempted PFR procedures were considered technically successful. Procedural details, type of MVP device implanted, associated interventions, complications, and re-interventions are summarized in **Table 2**, **Supplemental Table 2 and 3**. Additional interventions at the time of the PFR procedure were performed in 7 (41%) patients: the PDA was stented in 5 (29%) and a septal communication was created or enlarged in 2 (12%). The most frequent acute procedural complication was distal PFR migration (n=5, 29%), followed by access-related complications (n=4, 23%). One patient with L-looped ventricle experienced transient complete heart block requiring temporary pacemaker placement. In the initial peri-procedure period, a mild transient hemolysis was observed in 59% (n=10) of the patients. After the PFR procedure, the median mechanical ventilation duration was 116 hours (IQR 47-192). The vasoactive inotropic score decreased significantly from a median of 8 (IQR 0-11) at 6h post-procedure to a median of 2 (IQR 0-9, p=0.025) at 24h.

**Table 2:** Procedural details, acute and follow-up complications, target surgery, and patient-level outcomes.

| **Variable** | **Total**<br>**(n=17)** |
| --- | --- |
| Additional interventions during PFR procedure |  |
| PDA stenting | 5 (29.4) |
| Atrial septum intervention | 2 (11.7) |
| Procedural complications |  |
| Pulse loss (resolved) | 3 (17.6) |
| Femoral vein thrombosis (resolved) | 1 (5.9) |
| Acute PFR migration | 5 (29.4) |
| Partial | 1 (5.9) |
| Complete | 4 (23.5) |
| Pulmonary artery wire-injury | 1 (5.9) |
| CHB (resolved) | 1 (5.9) |
| PFR thrombosis | 1 (5.9) |
| Follow-up complications |  |
| Late PFR migration | 7 (41.1) |
| Partial | 2 (11.7) |
| Complete | 5 (49.3) |
| PFR thrombosis | 1 (5.9) |
| PFR collapse | 2 (11.7) |
| Vasoactive-inotropic support |  |
| At 6 hours | 8 (0-11) |
| At 24 hours | 2 (0-9) |
| Invasive MV duration, hours | 116 (47-192) |
| Additional interventions in follow-up | 7 (41.1) |
| PFR adjustment/revision | 4 (23.5) |
| Others | 6 (35.2) |
| PDA stenting | 5 (29.4) |
| Atrial septum intervention | 6 (35.2) |
| Coarctation stenting | 1 (5.9) |
| Balloon aortic valvuloplasty | 1 (5.9) |
| Target surgery, n (%) | 10 (58.8) |
| Age, days, median (IQR) | 96 (41-138) |
| Weight, kg, median (IQR) | 4.64 (3.20-5.42) |
| Weight gain since the PFR procedure, kg | 1.39 (0.62-2.88) |
| Target surgery details (n=10) |  |
| Norwood Stage-1 | 7 (70.0) |
| Complete repair | 2 (20.0) |
| Biventricular repair | 1 (10.0) |
| Length of index hospitalization after PFR procedure<br>(to discharge or death), days | 80 (18-150) |
| All-cause Mortality | 6 (35.2) |
| Before target surgery | 4 (23.5) |
| After target surgery | 2 (11.8) |

Continuous data are presented as median (Inter Quartile Range-IQR= 25% and 75%), categorical data are presented as frequency (%). Abbreviations: CHB: complete heart block; IQR: inter-quartile range; Kg: kilogram; MV: mechanical ventilation; PDA: patent ductus arteriosus; PFR: pulmonary flow restrictor.

### PFR-related complications

Acute and late migration of the PFR devices was the most common complication (**Table 2**, **Supplemental Table 3**). Acute migration was observed in 5 (29%) patients, all on the right side: complete in 4, partial in 1. Late migration was observed in 7 (41%) patients: 2 on the right side, 4 on the left, and 1 bilaterally. In 4 patients with unprotected RUPA, an additional device was implanted: in 2 at the time of the first procedure, and in the other 2 during follow-up. One completely migrated left PA device was replaced during a follow-up catheterization.

In addition, significant intra-device thrombosis of one of the PFR devices was observed in 2 (12%) patients. In one of these patients, the left PA device thrombosed immediately after release and was replaced during the same procedure. In the other patient, the thrombosed left PA device was identified at a follow-up catheterization at 3 months, and an intra-device stent implantation was required to re-establish adequate flow to the distal vessels.

Finally, in 2 (12%) patients in whom the PFR devices were implanted using the right upper PA technique, the devices were found partially collapsed at 3.4 and 3.6 months after the implantation. In one of these patients, a second PFR was implanted next to the fractured one because percutaneous retrieval was not possible. The second patient underwent the target surgery shortly thereafter as planned.

### Follow-up hemodynamic data, target surgery, and patient-level outcomes

Follow-up hemodynamic data are presented in **Supplemental Table 4**. Among the 17 patients, 11 (65%) underwent a follow-up hemodynamic study at a median time of 2.3 months (IQR 1.0-4.9). The median mean PA pressure distal to the PFR was 18 mmHg (IQR 14-26), the median mean LA pressure was 10 mmHg (IQR 8-16), the median trans-pulmonary gradient was 10 mmHg (IQR 7-12), and the median pulmonary vascular resistance (PVR) was 2.6 iWU (IQR 1.2-3.7). In patients with complete migration of the PFR, the mean pressure measured in the exposed upper PAs ranged from 24 to 50 mmHg. Five patients had significant LA hypertension, with a median LA pressure of 16 mmHg (IQR 16-21), and all of them had significantly elevated PVR compared to those with normal LA pressure (median 3.7 iWU [IQR 3.2-4.5] vs median 1.5 iWU [IQR 1.0-2.3], respectively; p=0.004). All 5 patients had LA hypertension that was secondary to a highly restrictive PFO, which was consequently addressed.

In total, 8 patients (47%) underwent an additional non-PFR related transcatheter procedure before the target surgery. Enlargement or creation of an atrial communication was performed in 6 (35.2%) patients, placement of a PDA stent in 5 (29%), stent of a retrograde coarctation in 1 (6%), and balloon aortic valvuloplasty in 1 (6%). One patient (6%) underwent an unplanned surgical intervention for removal of an atrial stent for refractory atrial arrhythmia, with PFRs left in place.

Patient-level outcomes are detailed in **Figure 2, Table 2,** and **Supplemental Table 2**. At a median follow-up of 5.3 months (IQR 1.9-9.6), 13 (76%) patients were either successfully bridged to surgery (n=10, 59%) or waiting surgery (n=3, 17%). Surgery was performed after a median time of 2.6 months (IQR 1.2-3.4) since the PFR procedure. The median age and weight at target surgery were 3.2 months (IQR 1.4-4.6) and 4.6 kilograms (IQR 3.2-5.4), respectively. All patients gained weight since the PFR procedure, with a median weight gain of 1.4 kilograms (IQR 0.6-2.9, p=0.009, **Figure 3a**). Except for one patient, who had Cornelia De Lange syndrome, all weighed ≥3 kilograms at the time of surgery. Measurements of PA showed adequate growth over time, from an initial median Z score of 0.1 (IQR −0.6-1.0) to a final Z score of 1.2 (0.9-1.8) at the pre-surgical catheterization (p=0.003; **Figure 3b**). During the target surgery, all MVP devices were easily removed without complications or the need for arterioplasty. Generally, the devices were removed with gentle traction and brief electrocautery to the distal stent portion where it was attached to the pulmonary artery.

**Figure 2.**
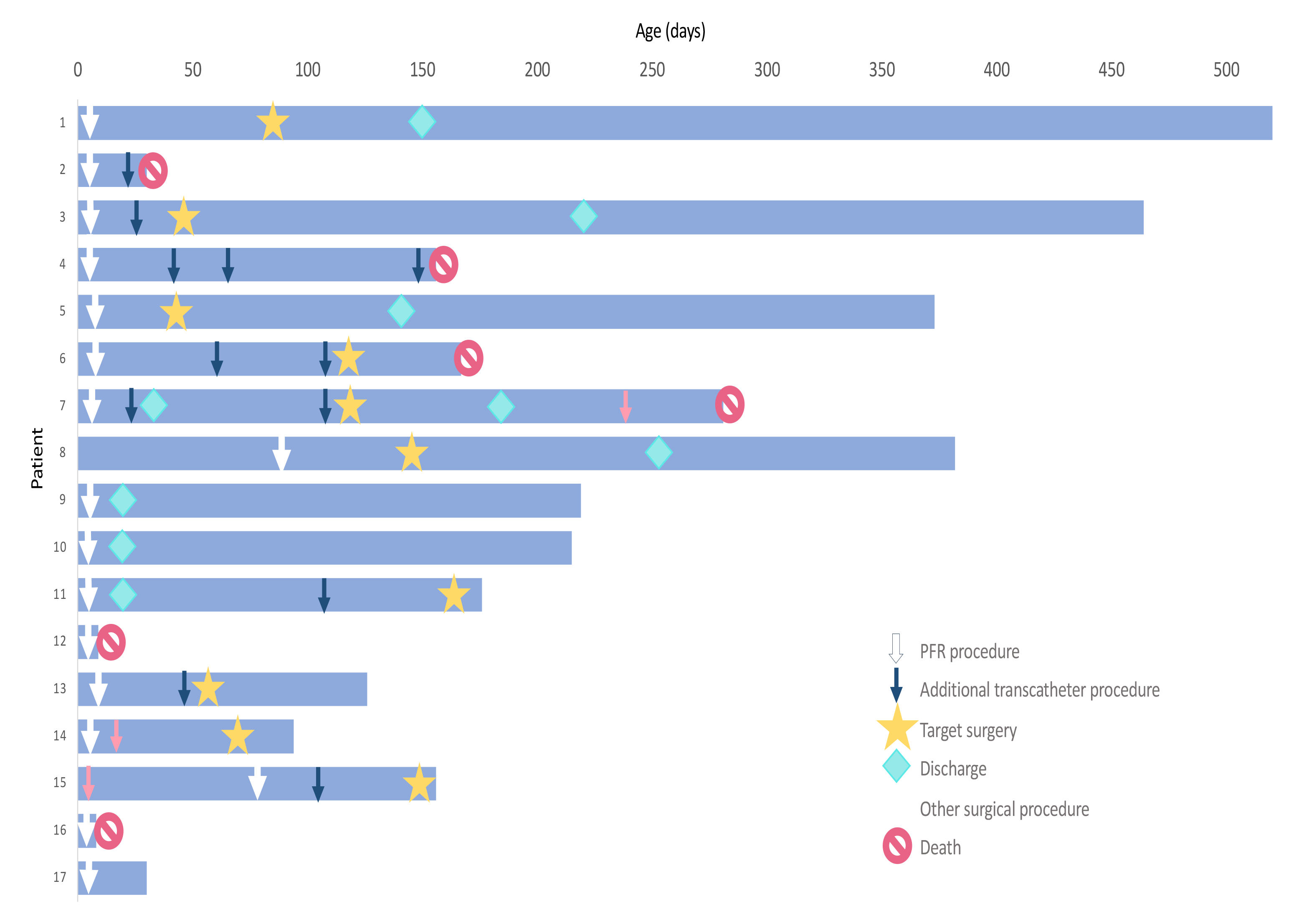
Summary of patient-level hospital courses and outcomes. PFR procedure is defined as the cardiac catheterization procedure where PFR was first implanted.

**Figure 3.**
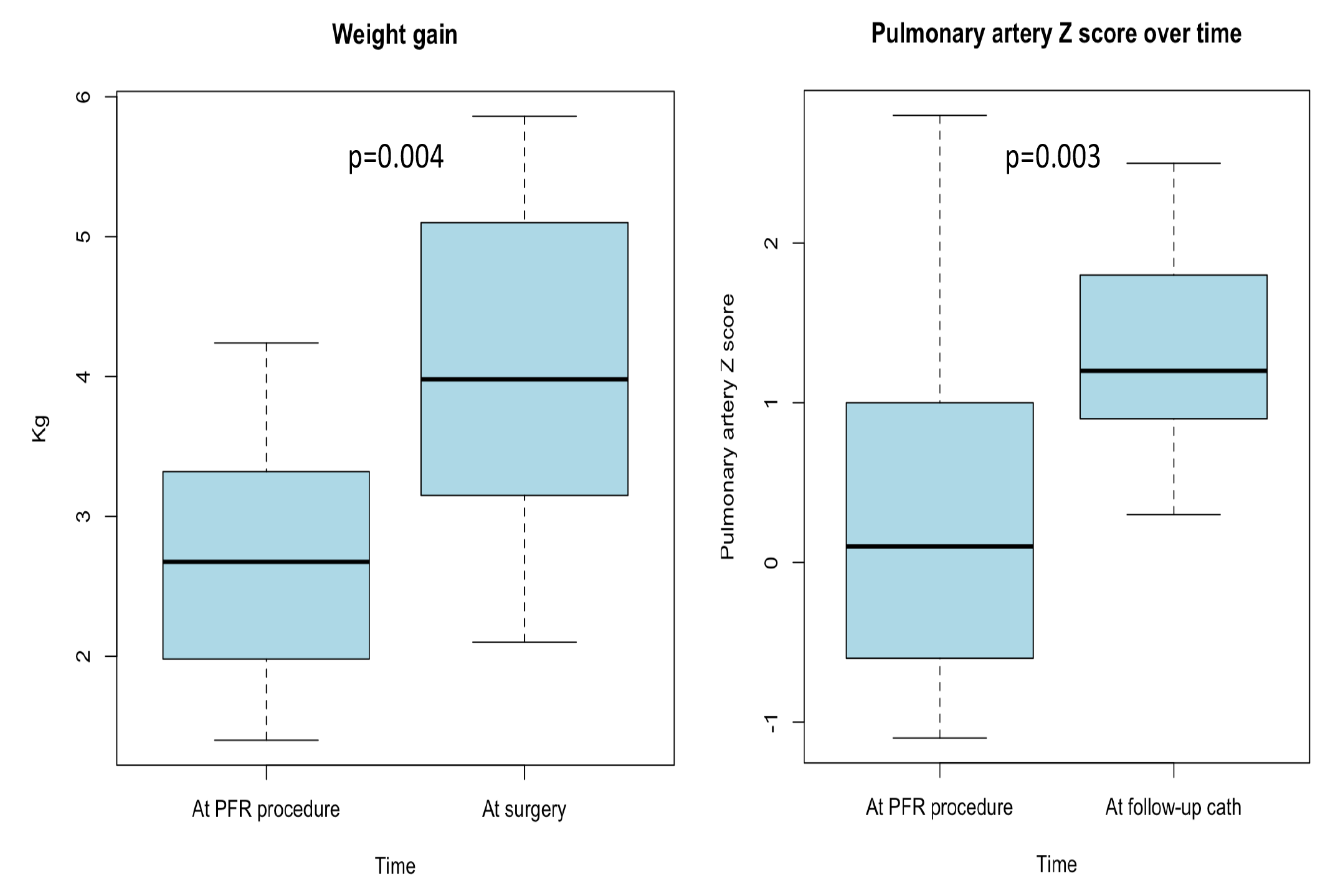
**Changes in weight and pulmonary artery diameter Z-scores over time.**

In the PFR cohort, 9 (53%) patients were discharged home either prior to (n=4) or following (n=5) the target surgery. Four patients died before target surgery and 2 after, for an overall all-cause mortality rate of 35%. Among the 4 deceased patients before target surgery, 2 were HLHS with both LV to coronary fistulae and IAS or severely restrictive PFO, 1 experienced complications following an emergent non-cardiac surgery on the first day of life, and 1 had a severe airway-related adverse event in the ICU. The estimated survival probabilities at 3 and 6 months were 82% (95% CI 65-98%) and 66% (95% CI 45-96%), respectively (**Figure 4**). The median length of hospitalization was 80 days (IQR 19-150).

**Figure 4.**
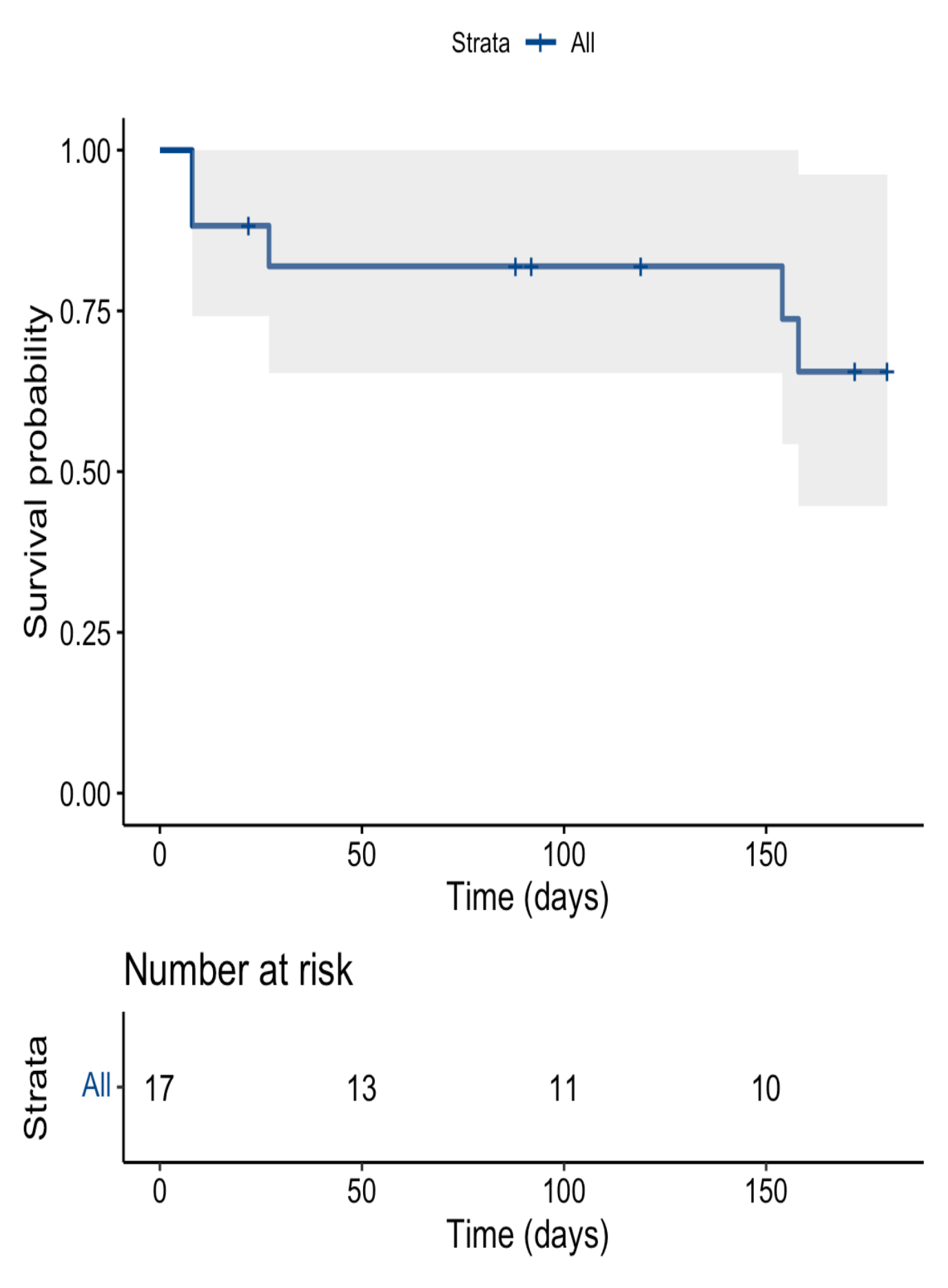
Kaplan Meier curve estimating the survival probability at 6 months after PFR procedure. The estimated survival probability at 6 months was 66% (95% CI 45-96%).

### Comparison between high-risk SV-PFR cohort and hybrid Stage 1 historical cohort

Between January 2012 and March 2023, 23 high-risk SV neonates underwent a hybrid Stage 1 palliation. Among the PFR cohort, 13 patients had SV physiology and a comparable high-risk profile (**Table 3**). Comparison of demographic and clinical characteristics showed no significant differences between the two cohorts, except for lower age at intervention in the PFR compared to the hybrid Stage 1 cohort (3 days, [IQR 2-4] vs 7 days [IQR 4-13], p=0.019). Additionally, patients in the PFR cohort had a higher frequency of IAS/severely restrictive PFO compared to the hybrid cohort (69% vs 17%, p=0.003). The estimated overall survival probabilities at 3 and 6 months were 76% (95% CI 56-98) and 46% (95% CI 21-98%), respectively, for the PFR cohort, 52% (95% CI 35-77%) and 30% (95% CI 16-56%), respectively, for the hybrid Stage 1 cohort. (**Figure 5**). After adjusting for age, weight, and presence of IAS, severely restrictive PFO or LV to coronary fistulae, the PFR cohort had a significantly lower 6-month all cause-mortality risk compared to the hybrid cohort (adjusted HR=0.30 [95% CI 0.10-0.93], **Figure 5, Table 4**). The same model showed no significant association at 3 months (**Supplemental Table 5**). The length of hospitalization did not significantly differ between the two cohorts (PFR: 80 days [IQR 22-148], hybrid: 42 days [IQR 6- 109]; p=0.217).

**Figure 5.**
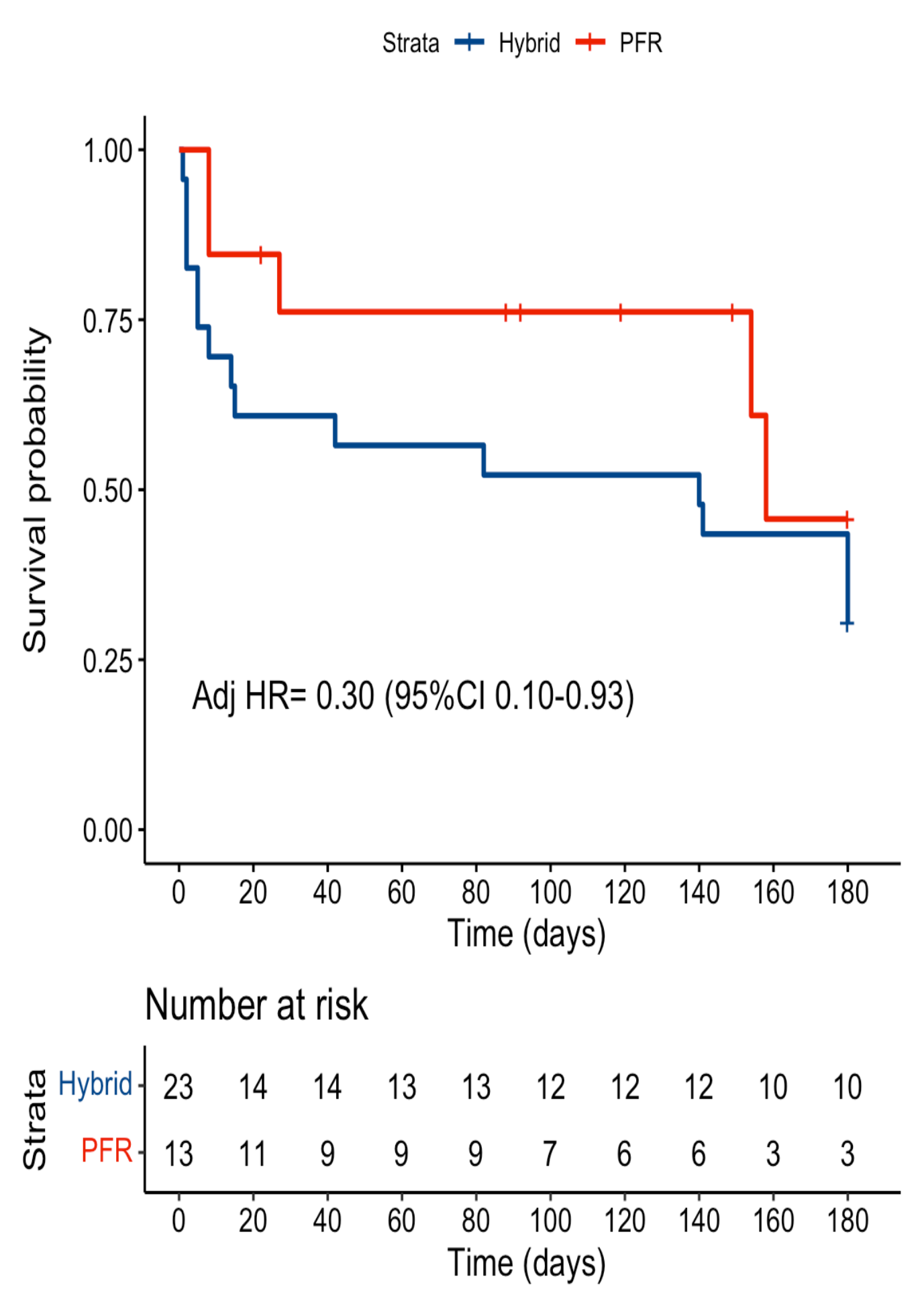
Kaplan Meier curves estimating survival probabilities at 6 months for the high-risk single ventricle PFR sub-cohort with the hybrid Stage 1 historical cohort. Estimated survival probabilities at 6 months were 46% (95% CI 21-98%) for the PFR cohort and 30% (95% CI 16-56%) for the hybrid cohort. When adjusting for age, weight, presence of IAS/severely restrictive PFO or LV to coronary fistula, the PFR cohort had a significantly lower 6-month mortality risk than the hybrid cohort (adjusted HR=0.30 [95% CI 0.10-0.93]).

**Table 3.** Demographic, anatomic, and clinical characteristic according to palliation strategy.

| Variable | High-risk SV patients undergoing PFR palliation (n=13) | High-risk SV patients undergoing hybrid palliation (n=23) | p value |
| --- | --- | --- | --- |
| Age at PFR procedure, days | 3 (2-4) | 7 (4-13) | **0.019** |
| Weight at PFR procedure, kg | 2.51 (2.20-3.20) | 2.50 (1.96-3.10) | 1.000 |
| Gestational age, weeks | 37 (34-38) | 36 (33-39) | 0.960 |
| Gender, male | 8 (61.5) | 13 (56.5) | 1.000 |
| Race |  |  |  |
| White | 6 (46.2) | 13 (56.5) | 0.075# |
| Black/African American | 2 (15.4) | 2 (8.7) |  |
| Others | 3 (23.0) | 2 (8.7) |  |
| Not available | 2 (15.4) | 6 (26.1) |  |
| Cardiac anatomic diagnosis |  |  |  |
| HLHS | 10 (76.9) | 17 (73.9) | 0.175# |
| DORV, hypoplastic LV | 2 (15.4) | 0 (0.0) |  |
| DILV | 0 (0.0) | 1 (4.3) |  |
| Unbalanced CAVC | 0 (0.0) | 3 (13.0) |  |
| Truncus, hypoplastic LV | 0 (0.0) | 1 (4.4) |  |
| LV aneurysm due to prenatal myocardial infarct | 1 (7.7) | 0 (0.0) |  |
| Dysplastic mitral valve with LVOT obstruction | 0 (0.0) | 1 (4.4) |  |
| Other anatomic risk factors |  |  |  |
| Intact atrial septum or severely restrictive PFO | 9 (69.2) | 4 (17.4) | **0.003#** |
| Left ventricle to coronary artery fistulae | 4 (30.7) | 4 (17.4) | 0.422# |
| Prematurity (<37 weeks of gestation), n (%) | 7 (53.8) | 12 (52.2) | 1.000 |
| Genetic syndrome and/or other congenital anomalies, n (%) | 4 (30.7) | 5 (21.7) | 0.693# |
| Length of hospitalization post-procedure, days (IQR) | 80 (22-148) | 42 (7-96) | 0.217 |

**Table 4.**
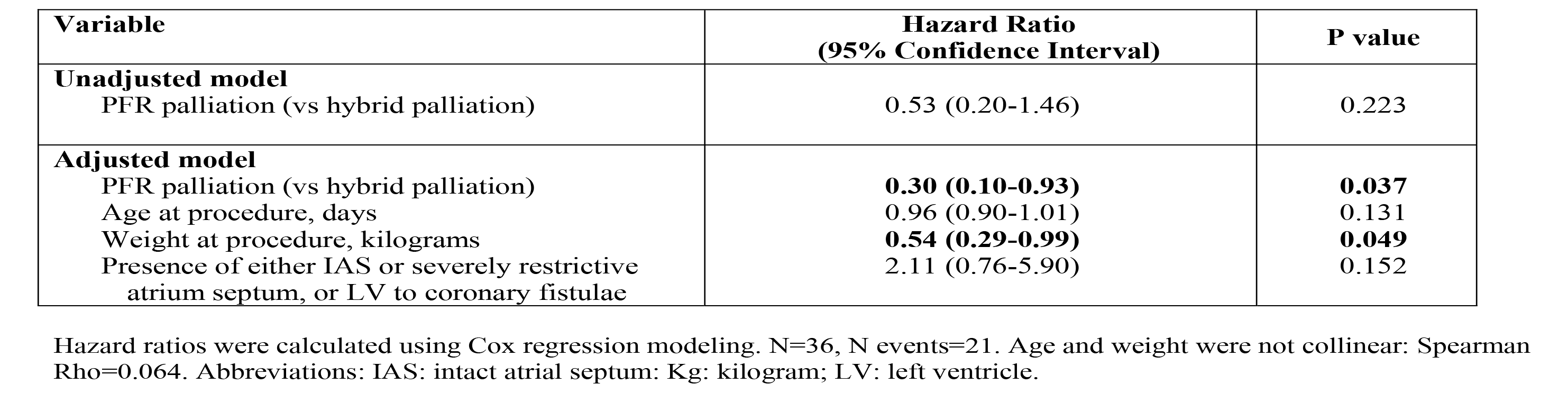
Cox regression model investigating the association between type of palliation and survival at 6 months.

## Discussion

This study demonstrated that transcatheter PFR-based palliation is feasible, safe, and represents an effective strategy for bridging high-risk neonates with CHD and pulmonary overcirculation to surgery later in life, while allowing for clinical stabilization and significant somatic growth. All procedures were technically successful, with a low incidence of severe adverse events despite the patients’ frailty. Most of the patients (76%) were either successfully bridged to target surgery or are hemodynamically stable while awaiting surgery. The PFR devices proved to be easily retrievable - both during transcatheter intervention and at the time of surgical operation - without complications or the need for arterioplasty. Moreover, PAs showed an adequate growth over time, which is particularly important for SV patients. Pre-surgical mortality was not procedure-related and was observed in patients with the increased mortality risk at baseline, such as those with SV and IAS/severely restrictive PFO or LV to coronary artery fistulae. Additionally, we showed that high-risk SV patients palliated with PFR had a decreased risk of short-term all-cause mortality compared to our historical cohort of high-risk SV patients palliated with a hybrid palliation strategy.

Various pre-clinical and clinical attempts at transcatheter regulation of PBF have been previously reported with varying degrees of success^13, 14^. In 2005, Boucek reported a case series of infants with HLHS palliated with a total transcatheter Stage 1 procedure as a bridge to heart transplant^15^. However, these techniques have not gained broad popularity due to the need for large delivery catheters to deploy the PFRs and challenges associated with their surgical removal. More recently, the encouraging results reported by Khan^11^ on the use of custom-made fenestrated MVP devices as PFR in a swine model led to a renewed interest in attempting to limit the PBF with a catheter-based approach^10^. To date, four clinical papers, including two small case series and two case reports^6–9^, have been published on this topic. Overall, the reported patients were mostly standard-risk surgical patients with SV physiology, and the success rates varied across reports. In the study by Schranz^6^, out of 6 newborns with SV physiology who underwent a total transcatheter Stage 1 palliation, 4 eventually underwent a comprehensive Stage 2, one a BiV repair, and one a transplant. Nageotte’s case series^9^ also reported 6 newborns who underwent a PFR-based palliation, all weighing >2.9 kg. Of these, 3 were bridged to target surgery (comprehensive Stage 2, complete repair, and BiV repair, respectively), one was converted to a Norwood Stage 1 shortly thereafter, and 2 had adverse outcomes. To the best of our knowledge, our cohort represents the largest reported series to date, and the first comprised mostly of high-risk surgical candidates. In fact, many of our patients had several universally recognized risk factors for mortality and adverse events post-surgery, including SV physiology, prematurity, low birth weight, genetic syndrome or congenital anomalies, other major comorbidities, preoperative mechanical ventilation, and shock^16, 17^. Furthermore, many of the SV patients, who would have otherwise undergone the highest risk neonatal cardiac surgical procedure - the Norwood Stage 1 palliation^18–23^ - had anatomical features associated with adverse outcomes, such as IAS or highly restrictive PFO^24–27^, LV to coronary fistulae^28, 29^, preoperative systemic AVVR, and SV dysfunction^18, 29, 30^. For these high-risk patients, PFR-based palliation provided a feasible and less invasive strategy to bridge the patients to a later surgery or transplant, once hemodynamic stability had been achieved, prematurity-related comorbidities had improved, and weight had been significantly optimized.

As expected, given the predominance of high-risk patients, the pre-target surgery mortality rate was not negligible (24%). However, these patients had associated conditions such as a genetic syndrome or non-cardiac congenital anomaly requiring major non-cardiac surgery, a major non-cardiac adverse event, LV to coronary fistulae with IAS, or a severely restrictive PFO. As expected, LV to coronary fistulae are almost exclusively seen in the mitral stenosis/aortic atresia HLHS variant^31^, and are associated with a postoperative mortality rate as high as 50%^28^. Similarly, patients with HLHS and IAS or a highly restrictive PFO are known to be at risk of poor outcome, with a reported preoperative mortality of 21%^27^ and an overall 1-month mortality rate ranging from 52% to 62%^25, 27^. Of the 6 deceased patients in our cohort, three had both anatomical features, and thus a higher baseline mortality risk.

To better investigate outcomes, we compared our sub cohort of high-risk SV PFR patients with a similar historical cohort who underwent a hybrid Stage 1 procedure at our institution. Originally developed to avoid hypothermic cardiopulmonary bypass/circulatory arrest and its associated hemodynamic derangements in high-risk patients^32, 33^, the hybrid palliation was shown to be an effective alternative to palliate high-risk neonates. However, previous studies have shown that in-hospital and interstage mortalities did not differ significantly from patients with similar risk profiles who underwent a traditional Norwood Stage 1 procedure^5, 34–36^. In our analysis, after adjusting for potential confounding factors, the PFR-based procedure was found to be associated with a significantly lower risk of mortality at 6 months compared to the hybrid cohort. While larger, multicenter studies are needed to confirm these findings, we believe that they represent an important preliminary result, indicating that this procedure may offer real benefits to high-risk neonates, particularly those who have limited surgical options due to a prohibitive risk profile.

Although our preliminary results are promising, there are technical limitations that must be acknowledged. As previously described^9^, there is a risk of persistent overcirculation, particularly during follow-up. Initially, despite using smaller fenestration in the MVP devices compared to previous reports^6–9^, we observed evidence of suboptimal PBF control. This led us to refine the fenestration technique further, eventually creating a 1-1.5 mm hole with a fine eye cauterizer. Additionally, as previously noted^8, 9^, the distal migration of PRF devices and the subsequent complete or partial exposure of the upper PAs, represents the true Achille’s heel of the procedure, with an incidence in our cohort of up to 70% (29% acute and 41% late). This phenomenon likely occurs due to the elastic and compliant nature of native PAs, whose systolic diameters change remarkably with the patient’s systemic blood pressure and volume state. Furthermore, the MVP devices do not appear to cause any substantial inflammatory response of the arterial wall, with minimal - if any - foreign body reaction, as demonstrated by the ease of retrievability of the devices even months after implantation. As a result, the PAs continue to grow around the devices, making an initially appropriately sized device undersized over time. Device migration also poses a risk of exposing the upper lobe arteries to unprotected PBF, which, in our experience, was often subclinical and discovered incidentally at follow-up catheterization. To mitigate the risk of embolization, the implanted devices were initially oversized compared to the manufacturer’s recommendations, but this approach had limited success. Subsequently, we found that the deployment of the device with its distal tip inside the take-off of the right upper PA could prevent any distal migration on the right side. Unfortunately, this technique could increase the mechanical stresses on the device frame that may eventually lead to fracture and partial collapse of device. In some patients, two devices were eventually implanted on the right side to protect the upper PAs, and we believe this two-device approach may represent the best strategy for protecting the right lung. However, neither of these techniques is feasible on the left side due to the short anatomical distance between the branch PA bifurcation and the take-off of the left upper PA. Nonetheless, we believe that the exposure of only the left upper PA to a higher than ideal pressure can be tolerated, particularly in those patients who will eventually undergo a BiV repair or a comprehensive Stage 2 within the first three to four months of life.

As with any new procedure, there is an initial learning curve that can lead to initial suboptimal results. Additionally, given the lack of long-term data, a closer follow-up must be strongly advocated. Indeed, in some cases, patients who were clinically asymptomatic were incidentally found to have migrated devices with unprotected segments of the lung, or significant LA hypertension and resultant pulmonary hypertension in the setting of a highly restrictive atrial communication. As a result, we recommend that all patients undergo an elective follow-up hemodynamic study, ideally at 6-10 weeks, regardless of their clinical status or echocardiographic findings.

It is important to note that this study has limitations, including small sample size and retrospective design. The patients included in the study were heterogeneous, which made the selection of an appropriate comparison cohort challenging. For this reason, we compared a subgroup of high-risk SV patients with a similar high-risk cohort who underwent a hybrid procedure, adjusting for multiple confounding variables. Because of the short follow-up time, we were only able to compare outcomes up to 3 and 6 months after the procedure. Ongoing follow-up will be necessary to investigate the interstage period and long-term outcomes. Despite this, our study represents the largest case series to date and provides new insights on this novel procedure and its outcomes.

In conclusion, we showed that transcatheter PFR-based palliation in neonates with CHD is feasible, technically safe, and an effective strategy for bridging to surgical palliation, complete repair, or transplant while allowing for clinical stabilization and significant somatic growth, especially in high-risk neonates with limited surgical options. Despite the implanted PFR devices, the PAs demonstrated adequate growth over time, and the devices were easily removed at the time of surgery without complications or need for arterioplasty. Mortality was not related to the procedure and was observed in patients with the highest expected baseline mortality risk. Although the results are promising, it is important to note that complications such as device migration may occur, and further research is needed to further optimize the PFR device, procedure, and its outcomes.

## Data Availability

Upon motivated request, data pertaining to this manuscript can be made available

## Nonstandard Abbreviations and Acronyms

AVVR: atrioventricular valve regurgitation
CHD: congenital heart disease
HLHS: Hypoplastic left heart syndrome
IAS: intact atrial septum
LA: left atrial
LV: left ventricle
MVP: Micro vascular plug
PA: pulmonary artery
PBF: pulmonary blood flow
PFO: patent foramen ovale
PFR: pulmonary flow restrictor
RUPA: right upper pulmonary artery
SV: single ventricle

## Acknowledgments

The authors thank Kai-ou Tang, MA, Medical Illustrator at Boston Children’s Hospital, Harvard Medical School, for her artistic contribution to Figure 1.

## Supplemental Material

Supplemental Methods

Tables S1-S5

